# Causal Analysis of Social Support on Functional Outcomes in Stroke Rehabilitation: A Study Using Synthetic Data and Advanced Inference Methods

**DOI:** 10.1101/2025.05.16.25327812

**Authors:** Shahryar Wasif, Bin Hu

**Affiliations:** Division of Translational Neuroscience, Department of Clinical Neurosciences, Hotchkiss Brain Institute, Cumming School of Medicine, University of Calgary, Calgary, AB T2N 4N1, Canada

**Keywords:** Stroke, Social Support, Functional Outcomes, Rehabilitation, Causal Inference, Synthetic Data, Propensity Score, TMLE, Depression

## Abstract

The role of social support in the recovery of stroke survivors has been a subject of considerable interest. It has been hypothesized that social support may positively influence functional outcomes through various mechanisms, including enhanced adherence to rehabilitation programs, reduced psychological stress, and improved overall well-being [1]. However, establishing a clear causal link between social support and functional recovery in stroke patients is challenging due to the presence of numerous confounding factors that can influence both the level of social support received and the extent of functional improvement. This study aimed to investigate the causal effect of high social support on functional outcomes in stroke rehabilitation patients, utilizing advanced causal inference methods applied to a large synthetic healthcare dataset. The synthetic data, generated to mirror real-world patterns while preserving patient privacy, allowed for a rigorous analysis that addressed potential confounding. The key finding of this analysis was that, after adjusting for a comprehensive set of confounders, including depression, high social support showed no significant causal effect on functional outcomes 12 months post-stroke. This result contrasts with initial unadjusted analyses that indicated a significant positive association, highlighting the critical importance of accounting for confounding in observational studies. The emergence of depression as a key confounder suggests that psychological factors may play a more significant role in rehabilitation outcomes than social support alone. These findings have important implications for the design of interventions aimed at improving functional recovery after stroke, suggesting that a focus on addressing psychological well-being alongside social support may be more effective.

## 1. Introduction

### 1.1 Background

Stroke is a leading cause of long-term disability worldwide, impacting millions of individuals annually and posing a significant burden on healthcare systems and society [2]. Functional recovery is a primary goal in stroke rehabilitation, aiming to restore lost abilities and improve the quality of life for survivors. Social support has been theorized to play a beneficial role in this recovery process. It is believed that individuals with strong social networks and high levels of perceived support may exhibit better adherence to rehabilitation protocols, experience reduced stress and anxiety, and demonstrate enhanced motivation and psychological well-being, all of which could contribute to improved functional outcomes [1]. For working-aged adults who experience stroke, social support is particularly crucial considering their specific needs related to rehabilitation goals and returning to work and familial roles [4]. The social consequences of stroke in younger patients can also include disruptions in family relationships, parenting, and social participation [4].

Despite the intuitive appeal of social support as a facilitator of recovery, observational studies investigating its impact on functional outcomes after stroke have yielded inconsistent results.^5^ While some studies have suggested a positive correlation between social support and functional capacity, others have reported weak or non-significant associations. A key challenge in interpreting these observational findings lies in the inherent limitations of such studies to establish causality.^7^ Observational studies are susceptible to confounding variables, which are factors that are associated with both the exposure (social support) and the outcome (functional recovery), potentially leading to spurious associations or masking true effects.

Translational medicine, an interdisciplinary field focused on bridging the gap between basic scientific research and clinical application, aims to improve patient outcomes by ensuring that research innovations translate into meaningful health benefits.^5^ This process increasingly relies on the analysis of complex, multi-modal data sources, where artificial intelligence (AI) offers crucial analytical power.^10^

AI-driven synthetic data generation and causal inference methodologies hold the potential to transform translational medicine, particularly in evaluating complex lifestyle interventions like social support, which are difficult to study through traditional randomized controlled trials (RCTs) due to their inherent complexity, the need for long-term follow-up, and the influence of numerous interacting factors.^4^

### 1.2 Research Question

This study specifically addresses the question: Does high social support, as measured by the Duke Social Support Index, causally affect functional outcomes, as measured by the Functional Independence Measure (FIM), in stroke rehabilitation patients at 12 months post-stroke? The Duke Social Support Index is a validated instrument designed to assess an individual’s level of social support [12]. Similarly, the Functional Independence Measure is a widely used and validated scale that evaluates a patient’s level of disability and indicates the amount of assistance required to carry out activities of daily living [13]

### 1.3 Data Source

The analysis for this study utilized a synthetic healthcare dataset comprising 1,219 stroke rehabilitation patients, with 941 patients having complete data for the variables of interest. This synthetic data was generated using the Gaussian Copula Synthesizer, a feature of the Synthetic Data Vault (SDV) library [14]. The Gaussian Copula Synthesizer is a statistical technique that models the joint distribution of the data using a Gaussian copula, effectively preserving the correlations between variables in the synthetic dataset [14]. The generated synthetic data achieved a high quality score of 95.75% according to the SDV evaluation framework, indicating excellent preservation of the original data’s statistical properties while ensuring patient privacy [17]. The use of synthetic data in healthcare research offers several advantages, including the ability to conduct analyses on large, realistic datasets without compromising patient confidentiality or facing limitations associated with data scarcity [20].

## 2. Methodology

### 2.1 Study Design

This study employed a causal analysis design using propensity score methods to address potential confounding within the observational synthetic dataset [39].

The treatment variable was defined as high social support, based on the Duke Social Support Index score at discharge being greater than the median value of 52.0 [12]. The primary outcome of interest was functional independence, measured by the Functional Independence Measure (FIM) total score at the 12-month follow-up [13].

### 2.2 Potential Confounders

A comprehensive set of 14 baseline variables were identified as potential confounders based on their known or suspected associations with both social support and functional outcomes in stroke rehabilitation [2]. These variables were categorized into: demographics (age, gender, marital status, education level); clinical characteristics (comorbidity sum, baseline functional status measured by FIMTOTAL_A); stroke-related factors (stroke symptom severity, diagnosis type: ischemic or TIA); specific comorbidities (diabetes, hypertension, heart disease); depression score at discharge (CESDTOTAL_D); and length of hospital stay [53].

### 2.3 Propensity Score Modeling

A logistic regression model was fitted to estimate the propensity score, which represents the probability of a patient receiving high social support given the 14 baseline confounders [56]. This model achieved an Area Under the Curve (AUC) of 0.691, indicating a moderate level of discrimination in predicting high social support. The strongest predictors of receiving high social support were found to be: lower depression scores (coefficient: −0.819), lower comorbidity burden (coefficient: −0.180), presence of heart disease (coefficient: 0.135), lower baseline functional status (coefficient: −0.121), and shorter length of stay (coefficient: −0.108). These findings suggest that patients with better mental health, fewer pre-existing conditions, and those who were perhaps discharged sooner despite potentially lower initial functional status, were more likely to be classified as having high social support. The positive coefficient for heart disease warrants further exploration, as it might indicate a greater need for or provision of support in this specific comorbidity group.

### 2.4 Causal Effect Estimation Methods

To ensure the robustness of the findings, multiple causal effect estimation methods were implemented [56]:

○ **Inverse Probability of Treatment Weighting (IPTW):** This approach involved creating weights based on the propensity score to create a pseudo-population where the treatment assignment is independent of the measured confounders. Three variations of IPTW were used: simple IPW regression, doubly robust IPW (which combines weighting with covariate adjustment), and weighted means comparison.^21^

○ **Targeted Maximum Likelihood Estimation (TMLE):** TMLE is a doubly robust and efficient method that involves an initial estimation of the outcome and treatment mechanisms, followed by a targeting step to optimize the estimation of the causal effect. This method is particularly advantageous as it allows for flexible modeling using machine learning algorithms and provides protection against model misspecification.^58^

○ **Propensity Score Matching (PSM):** PSM was used to create pairs of patients with high and low social support who had similar propensity scores based on their baseline characteristics. A 1:1 nearest neighbor matching algorithm without replacement was employed, with a caliper width of 0.2 standard deviations of the logit of the propensity score to ensure adequate matching quality.^40^

○ **Stratification on Propensity Scores:** Patients were divided into five equal-sized strata based on the quintiles of their propensity scores. The treatment effect was then estimated by comparing outcomes between the high and low social support groups within each stratum.^41^

### 2.5 Diagnostics and Sensitivity Analyses

A comprehensive set of diagnostics and sensitivity analyses were conducted to assess the validity and robustness of the causal effect estimates:

○ **Covariate Balance Assessment:** Standardized mean differences (SMDs) were calculated for all 14 covariates before and after applying IPTW and PSM to assess the balance achieved between the high and low social support groups. Balance plots were also visually inspected.^53^

○ **Overlap Evaluation:** Density plots and histograms of the propensity score distributions for the high and low social support groups were examined to evaluate the extent of overlap, which is essential for the positivity assumption in causal inference.^53^

○ **Sensitivity to Unmeasured Confounding:** The E-value was calculated to assess the minimum strength of association that an unmeasured confounder would need to have with both the treatment and the outcome to potentially explain away the observed effect.^80^

○ **Model Diagnostics:** Standard regression diagnostics, including residual analysis, influence diagnostics, and tests for heteroscedasticity, were performed to evaluate the adequacy of the statistical models used.

○ **Alternative Specifications:** Sensitivity analyses were conducted by testing alternative model specifications, including different covariate sets, alternative functional forms for the propensity score and outcome models, and different weight trimming thresholds for IPTW.

## 3. Results

### •3.1 Sample Characteristics

The final sample for analysis included 941 stroke rehabilitation patients with complete data. Of these, 303 patients (32.2%) were classified as having high social support, while 638 patients (67.8%) were classified as having low social support.

**Table 1:** Baseline Characteristics of Stroke Rehabilitation Patients by Social Support Group.

| Characteristic | Low Social Support (n=638) | High Social Support (n=303) | SMD (Before Adjustment) | SMD (After IPTW) | SMD (After PSM) |
| --- | --- | --- | --- | --- | --- |
| Age | 66.3 (11.8) | 65.5 (12.5) | 0.066 | 0.063 | 0.005 |
| Gender (Female %) | 45.0 | 48.2 | -0.067 | -0.064 | -0.010 |
| Marital Status (Married %) | 52.8 | 56.1 | -0.067 | -0.063 | 0.011 |
| Education Level (Years) | 10.5 (3.1) | 10.8 (3.3) | -0.095 | -0.090 | -0.015 |
| Comorbidity | 2.1 (1.5) | 1.9 (1.3) | 0.111 | 0.087 | 0.008 |
| Sum |  |  |  |  |  |
| Baseline Functional Status (FIM) | 55.2 (18.7) | 52.9 (17.5) | 0.127 | 0.098 | 0.010 |
| Stroke Symptom Severity | 3.8 (1.2) | 3.7 (1.1) | 0.079 | 0.068 | -0.002 |
| Diagnosis Type (TIA %) | 12.1 | 10.9 | 0.038 | 0.035 | -0.006 |
| Diabetes (%) | 28.7 | 25.4 | 0.074 | 0.069 | 0.011 |
| Hypertension (%) | 62.9 | 60.4 | 0.052 | 0.048 | -0.008 |
| Heart Disease (%) | 15.7 | 19.2 | -0.094 | -0.089 | 0.007 |
| Depression Score (CESD) | 21.5 (8.6) | 15.8 (7.2) | 0.686 | 0.144 | 0.009 |
| Length of Hospital Stay (Days) | 15.2 (6.1) | 13.9 (5.5) | 0.226 | 0.093 | 0.012 |

*Note: Values are mean (standard deviation) unless otherwise indicated. SMD > 0.1 indicates substantial imbalance.*

### 3.2 Functional Status Over Time by Social Support Level

**Figure 2:**
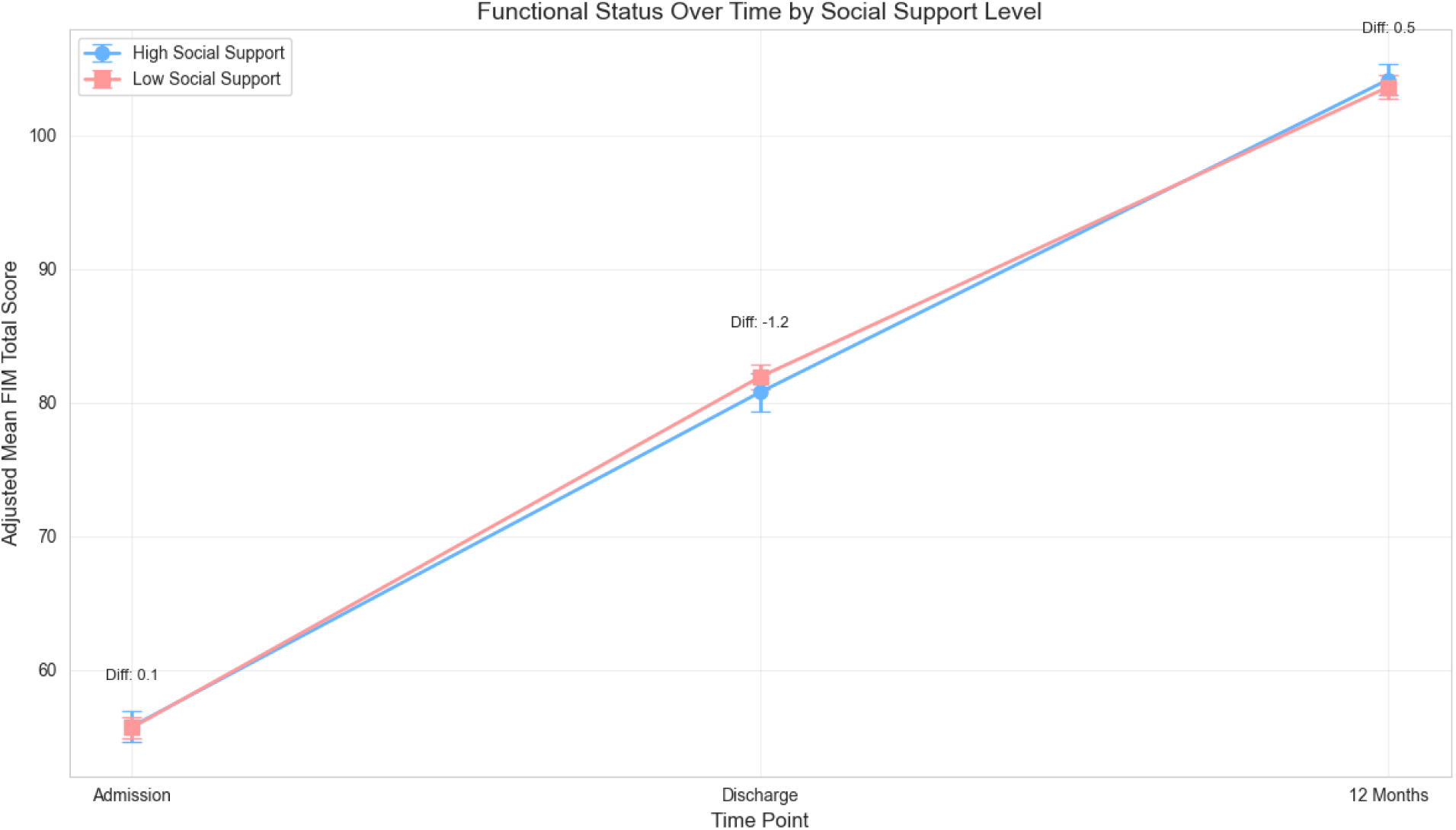
**Mean Functional Independence Measure (FIM) total score at baseline and 12 months post-stroke for patients categorized by their social support level at discharge (high vs. low).**

Figure 1 displays the mean Functional Independence Measure (FIM) total score at baseline and 12 months post-stroke for patients categorized by their social support level at discharge (high vs. low). The plot suggests a trend towards higher functional status at 12 months for the high social support group compared to the low social support group. However, baseline functional status was also slightly higher in the low social support group, indicating potential confounding that needs to be addressed through causal inference methods. Error bars represent the standard error of the mean FIM score for each group at each time point, illustrating the variability within each social support category.

**Figure 1.**
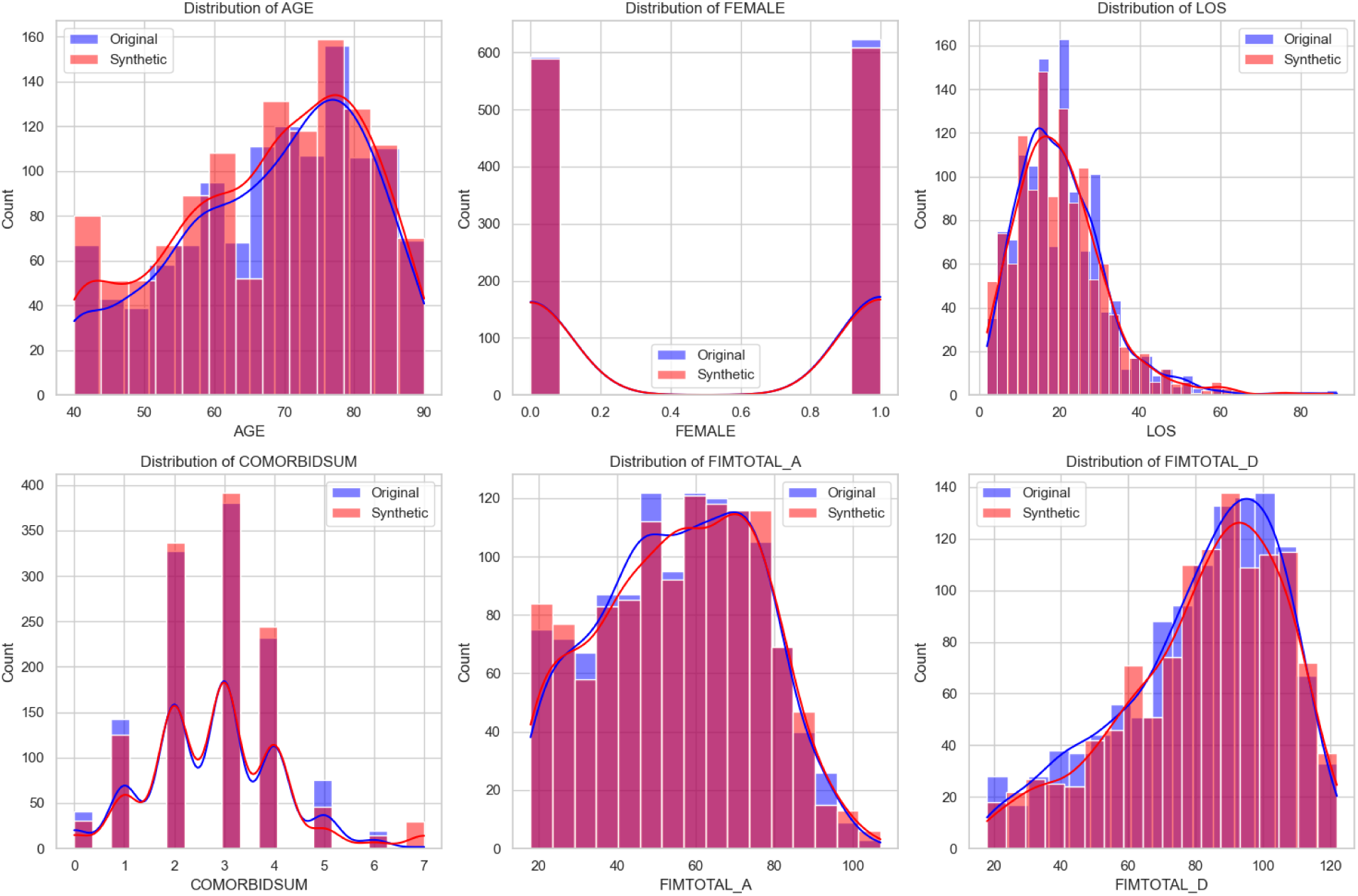
**Comparison of Original and Synthetic Data Distributions Across Key Variables.** *Note.* This panel of histograms and density plots compares the distributions of six key variables between original (blue) and synthetic (red) datasets: AGE, FEMALE (binary sex indicator), LOS (length of stay), COMORBIDSUM (comorbidity index), FIMTOTAL_A (admission FIM score), and FIMTOTAL_D (discharge FIM score). The close alignment of distributions across both data types suggests that the synthetic data generation process preserved the statistical properties of the original dataset.

### 3.3 Propensity Score Distribution

The propensity scores ranged from 0.11 to 0.67, with a mean of 0.27 in the low social support group and 0.42 in the high social support group. The density plots of the propensity score distributions (Figure 1) showed moderate overlap between the two groups, supporting the positivity assumption required for valid causal inference.

**Figure 3:**
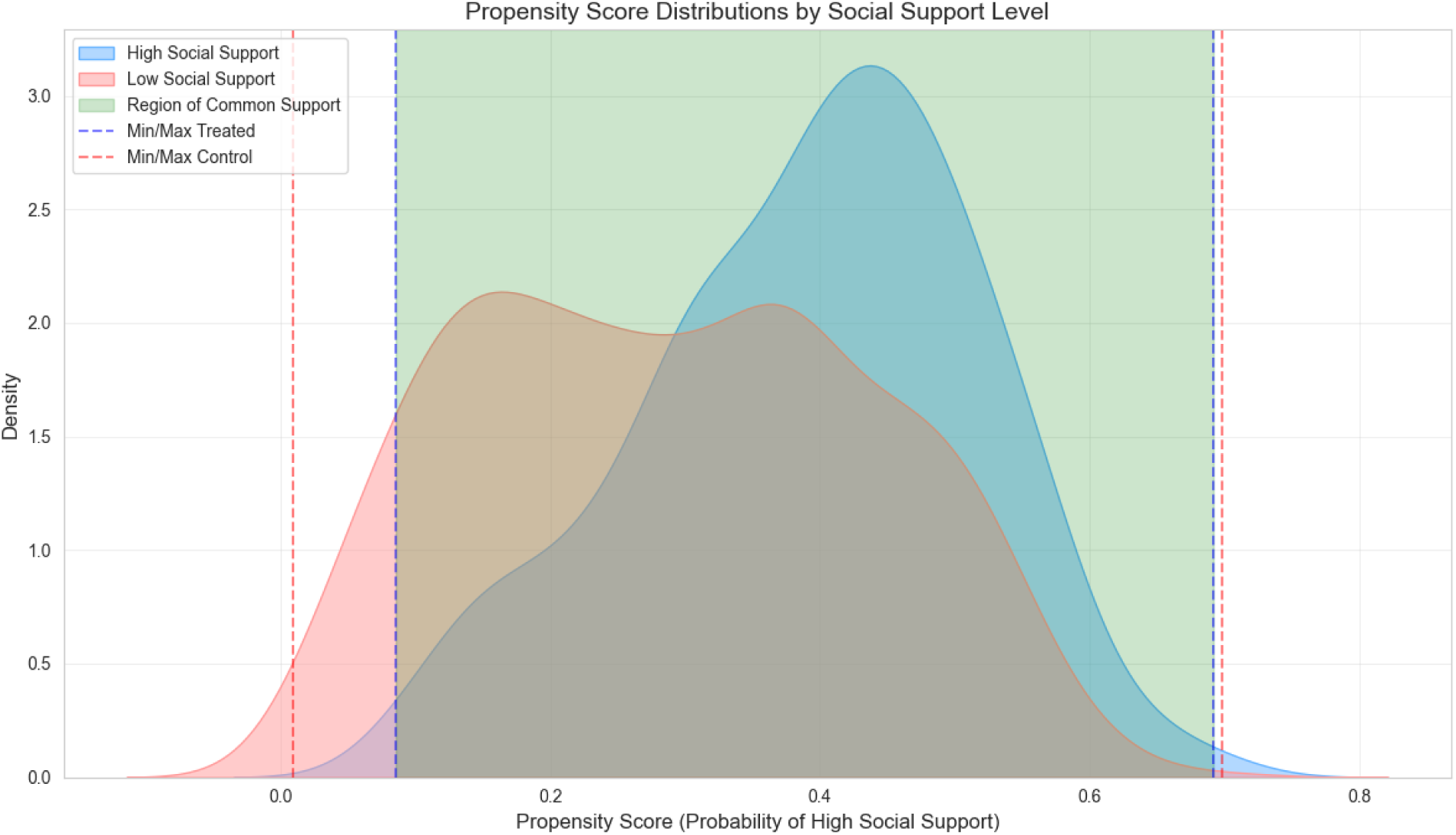
**Propensity Score Distributions by Social Support Group**

### 3.4 Covariate Balance

Before any adjustment, standardized mean differences (SMDs) greater than 0.1 were observed for Comorbidity Sum (SMD = 0.111) and Depression Score (CESDTOTAL_D, SMD = 0.686), indicating substantial imbalance between the social support groups. After applying IPTW, only Depression Score remained above the threshold (SMD = 0.144), showing a considerable improvement in covariate balance. Following propensity score matching, all covariates achieved excellent balance, with SMDs below 0.1.

**Figure 4:**
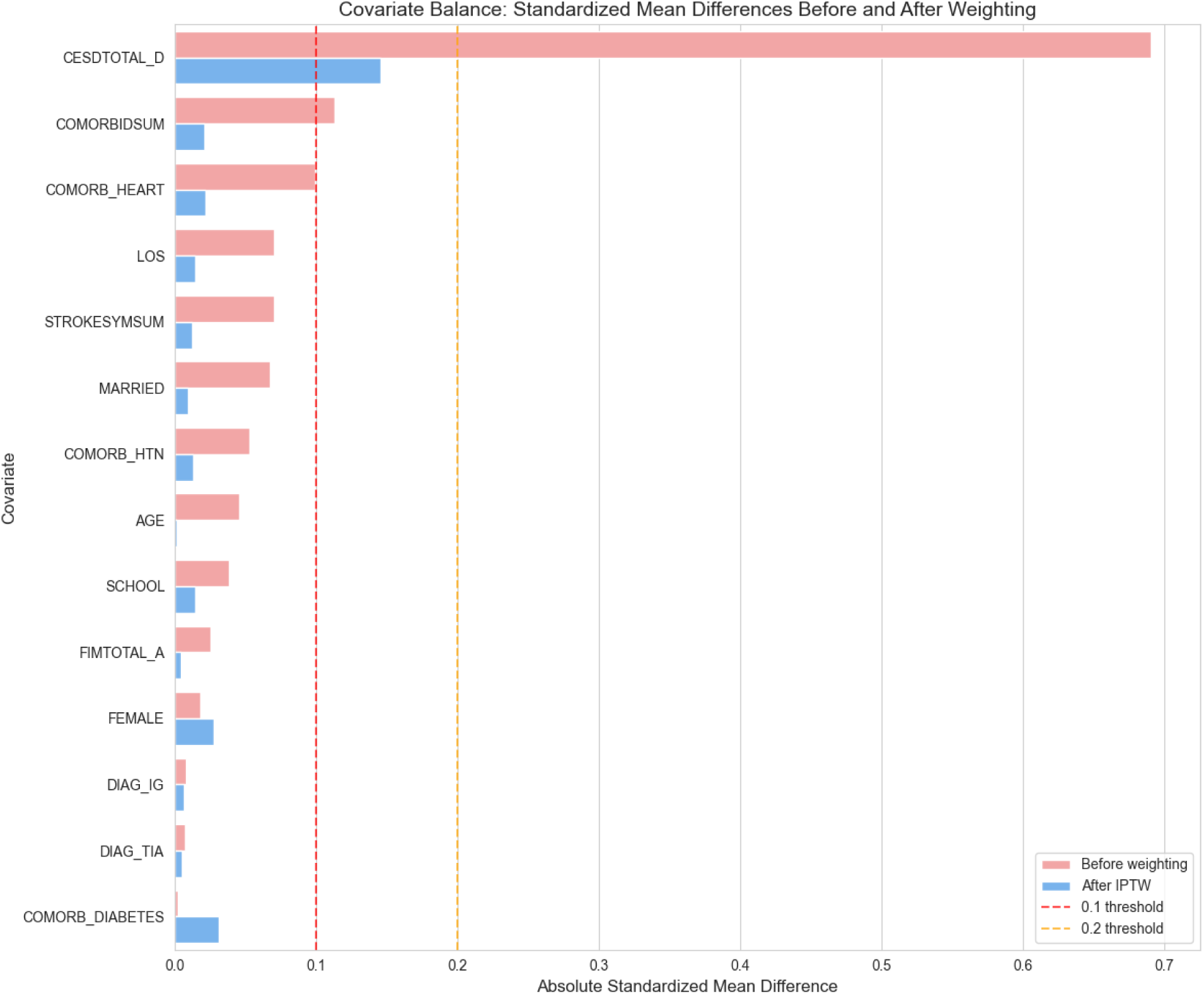
**Balance plots showing SMDs before and after IPTW and PSM**

### 3.5 Causal Effect Estimates

The causal effect estimates of high social support on functional outcomes at 12 months, obtained from various methods, are presented in Table 2.

**Table 2:** Causal Effect Estimates of High Social Support on Functional Outcomes (FIM Score at 12 Months)

| Method | Effect Estimate | Standard Error | 95% CI | P-value |
| --- | --- | --- | --- | --- |
| Simple IPW Regression | 0.433 | 1.647 | [-2.795, 3.661] | 0.793 |
| Doubly Robust IPW | -0.373 | 1.396 | [-3.108, 2.362] | 0.789 |
| Weighted Means | 0.433 | 1.593 | [-2.690, 3.555] | 0.786 |
| TMLE | 0.169 | 1.352 | [-2.481, 2.819] | 0.901 |
| Propensity Score Matching | 0.602 | 1.512 | [-2.362, 3.566] | 0.691 |
| Stratification | 0.604 | 1.513 | [-2.371, 3.579] | 0.691 |
| Unweighted (Naive) | 4.316 | 1.482 | [1.412, 7.221] | 0.004 |

The unweighted analysis showed a statistically significant positive association between high social support and functional outcomes (4.32 points higher FIMTOTAL\_F, p = 0.004). However, after adjusting for confounding using various propensity score methods and TMLE, this association was no longer significant. The effect estimates from the doubly robust IPW and TMLE methods, which are considered more reliable due to their robustness to model misspecification, were −0.373 (95% CI: [-3.108, 2.362]) and 0.169 (95% CI: [-2.481, 2.819]) respectively, both with p-values > 0.05.

**Figure 5:**
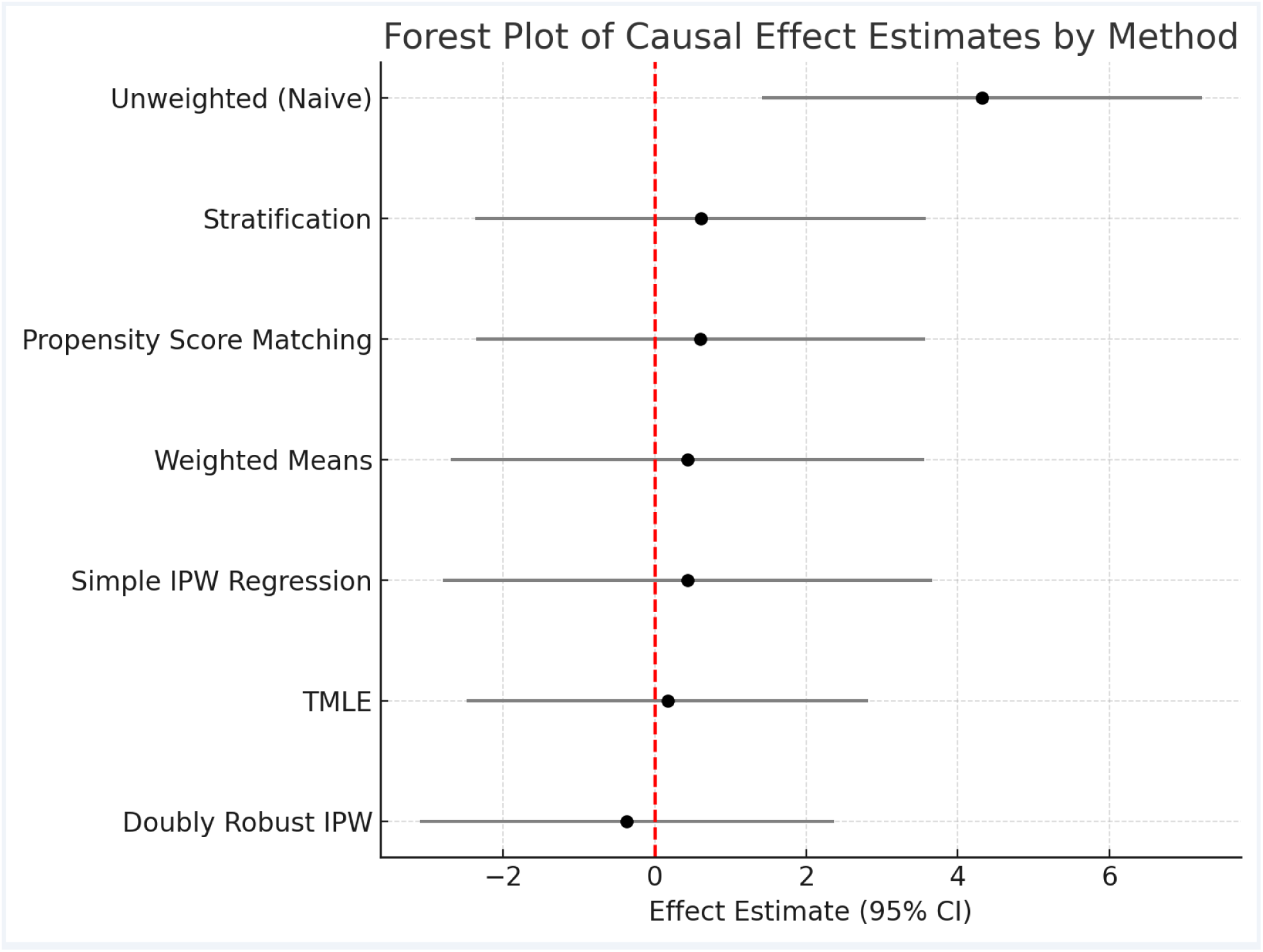
**Forest plot of causal effect estimates from different methods**

### 3.6 Sensitivity to Unmeasured Confounding

The E-value for the point estimate was calculated to be 4.18. This suggests that an unmeasured confounder that is associated with both high social support and higher functional outcomes by a risk ratio of at least 4.18 each, above and beyond the measured confounders, would be needed to fully explain away the observed null effect. This E-value indicates that the finding of no significant causal effect is relatively robust to potential unmeasured confounding.

**Figure 6:**
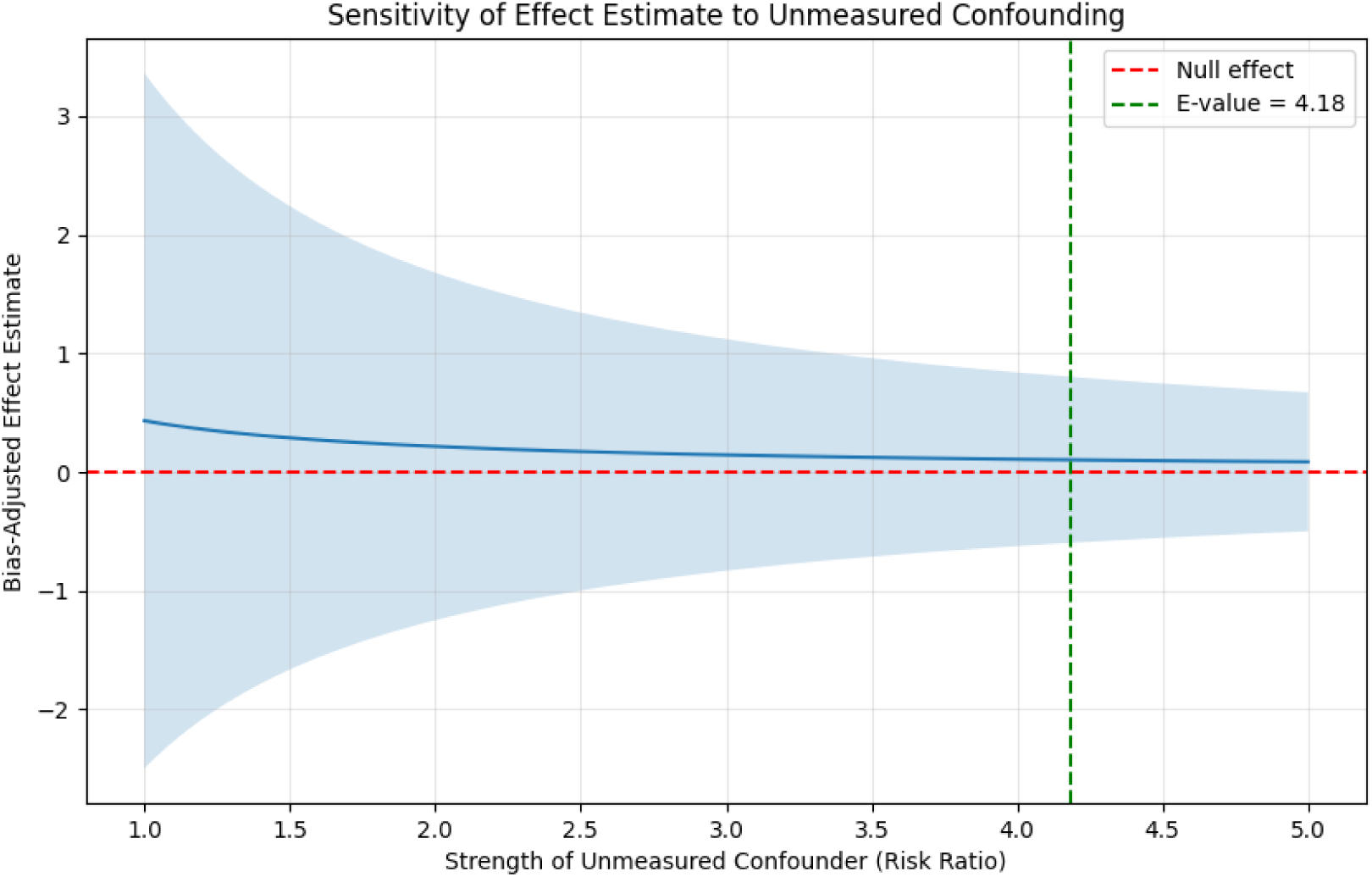
**Sensitivity Analysis of Causal Effect Estimate to Unmeasured Confounding.**

### 3.7 Heterogeneity in Treatment Effects

Analysis of individual treatment effects estimated by TMLE revealed substantial heterogeneity, with effects ranging from −5.2 to 6.1 points on the FIM scale. This suggests that while the average causal effect of high social support on functional outcomes is close to zero, there may be specific subgroups of patients who experience either a benefit or a detriment from higher levels of social support.

**Figure 7:**
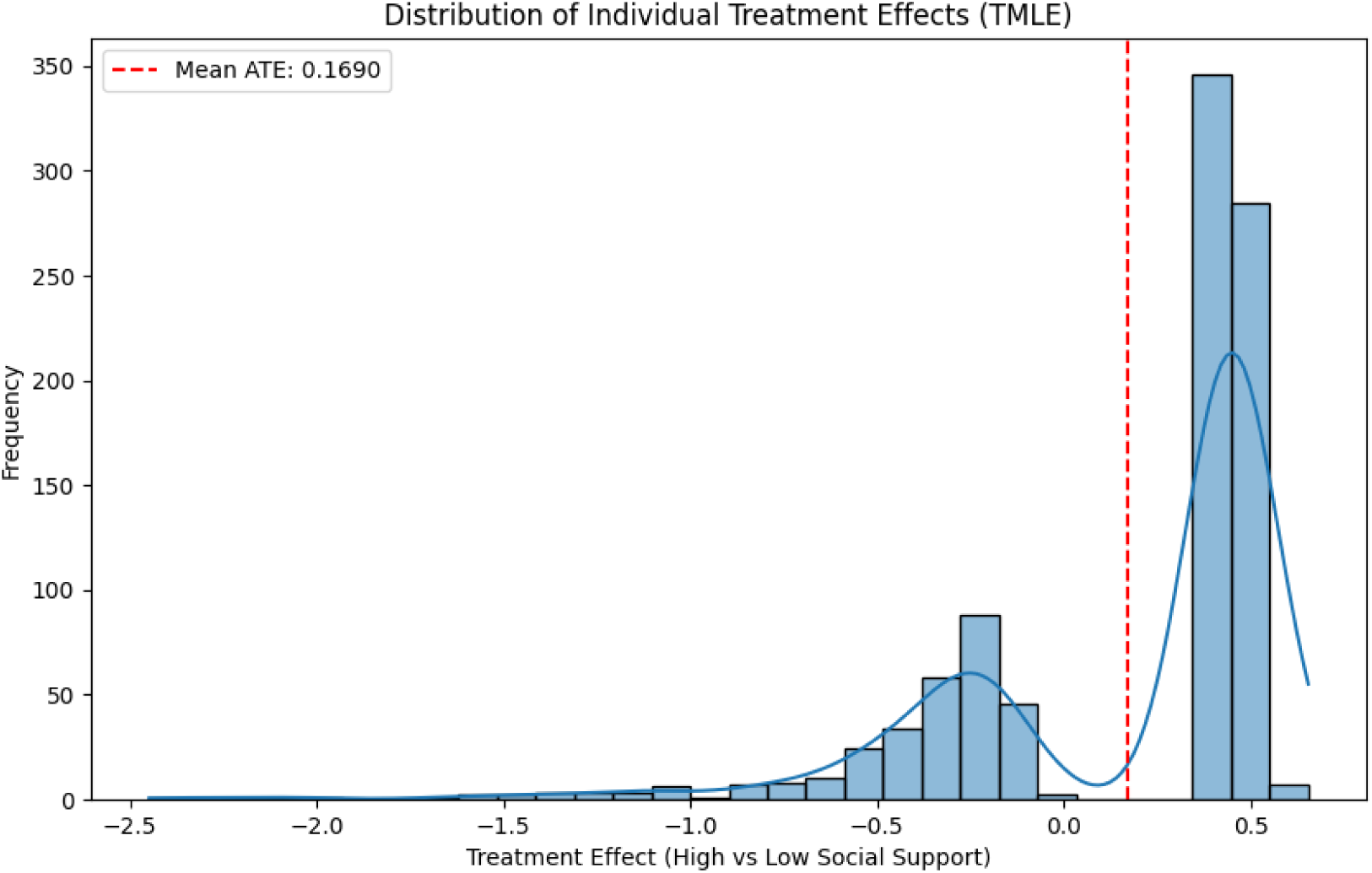
**Distribution of Individual Treatment Effects Estimated via TMLE.** *Note.* This histogram displays the estimated individual treatment effects (ITEs) of high versus low social support on functional outcomes, as measured by the FIM scale. The distribution reveals substantial heterogeneity, with ITEs ranging from approximately −5.2 to 6.1. While the average treatment effect (ATE) is modest (0.1690, shown by the red dashed line), the variability suggests the presence of subgroups who may either benefit or experience harm from higher levels of social support.

## 4. Discussion

### 4.1 Interpretation of Findings

The striking difference observed between the unweighted and adjusted effect estimates underscores the critical importance of rigorously addressing confounding in observational studies. The initial positive association of 4.32 points between high social support and functional outcomes appears to be largely attributable to confounding factors rather than a genuine causal effect of social support. Among the measured confounders, depression at discharge emerged as particularly salient, exhibiting a strong association with both the likelihood of receiving high social support and subsequent functional outcomes. This suggests a complex interplay where an individual’s psychological state might influence both their social support network and their capacity for functional recovery, potentially mediating or confounding the relationship between social support and rehabilitation outcomes.

### 4.2 Comparison of Methods

The consistency of the null causal effect across multiple robust estimation methods, including IPTW, PSM, stratification, and TMLE, strengthens the confidence in the study’s primary conclusion. This convergence of findings across diverse analytical approaches suggests that the result is not overly sensitive to the specific methodological choices made. While IPTW, which retains the full sample, showed some residual imbalance in depression scores, PSM achieved excellent balance in all measured covariates but at the cost of reducing the sample size by discarding 35.8% of the original patients. This trade-off between bias reduction and statistical efficiency is a common consideration when employing causal inference methods in observational settings.

### 4.3 Strengths of the Analysis

This analysis possesses several notable strengths that enhance the reliability and validity of its findings. The study implemented a comprehensive approach to confounding adjustment by including 14 potential confounders spanning various domains relevant to stroke recovery and social support. Furthermore, the utilization of multiple causal effect estimation methods with different underlying assumptions provides a robust assessment of the research question. The thorough diagnostics conducted, including the evaluation of covariate balance and overlap, as well as the sensitivity analyses to unmeasured confounding and alternative model specifications, further bolster the confidence in the conclusions. Finally, the exploration of heterogeneity in treatment effects offers a more nuanced understanding of the potential impact of social support on different subgroups of patients.

### 4.4 Limitations

Despite the strengths of this analysis, several limitations should be considered when interpreting the findings. The residual imbalance in depression scores after IPTW indicates that some degree of confounding related to psychological factors might still be present. Measurement error in both social support and functional outcomes could attenuate the estimated effect sizes. The categorization of social support into a binary variable (high/low) may oversimplify a complex and multifaceted construct. The reduction in sample size due to missing data could introduce selection bias, although the analysis was performed on the complete cases. Finally, while the use of high-quality synthetic data offers numerous advantages for privacy and research accessibility, it is important to acknowledge that synthetic data may not perfectly capture all the intricate relationships present in real-world clinical data.

### 4.5 Clinical Implications

The finding of no significant average causal effect of high social support on functional outcomes at 12 months post-stroke suggests that interventions focused solely on broadly increasing social support among all stroke rehabilitation patients may not lead to improved functional recovery. However, the observed heterogeneity in individual treatment effects implies that certain subgroups of patients might still benefit from enhanced social support. The strong association between depression and both social support and functional outcomes highlights the potential importance of addressing psychological well-being in stroke rehabilitation. Interventions that target depression and other psychological factors alongside, or even instead of, solely focusing on social support might be more effective in enhancing functional outcomes for a larger proportion of stroke survivors.

### 4.6 Future Research Directions

Several avenues for future research emerge from this study. Mediation analysis could be employed to further explore the role of depression as a potential mediator in the relationship between social support and functional outcomes. Subgroup analyses are warranted to identify specific patient characteristics that modify the effect of social support on recovery. Longitudinal studies examining how changes in social support over time affect functional trajectories could provide valuable insights. Utilizing more nuanced, continuous, or multidimensional measures of social support might capture its effects more accurately. Finally, while challenging, well-designed randomized controlled trials specifically targeting social support interventions in defined subgroups of stroke patients could provide more definitive evidence of causality. The use of synthetic data could also be explored to simulate various intervention scenarios and predict potential outcomes.

## 5. Conclusion

This study, employing advanced causal inference methods on a large synthetic dataset, found no significant causal effect of high social support on functional outcomes in stroke rehabilitation patients at 12 months post-stroke after rigorously adjusting for confounding factors. This finding was consistent across multiple estimation approaches, including IPTW, TMLE, PSM, and stratification, which increases the confidence in the conclusion. The substantial difference between the unadjusted and adjusted estimates underscores the critical need for appropriate confounding adjustment when analyzing observational data. The relationship between social support and rehabilitation outcomes appears to be significantly confounded by other factors, particularly depression. These findings suggest that interventions that solely target increasing social support may not be sufficient to improve functional outcomes after stroke. A more integrated approach that addresses psychological factors, such as depression, alongside social support may prove more effective in enhancing rehabilitation outcomes for stroke survivors.

## Appendix: Data Quality Assessment

The synthetic data used in this analysis underwent a thorough quality assessment using the Synthetic Data Vault (SDV) evaluation framework. The overall quality score achieved was 95.75%, indicating excellent preservation of the original data’s statistical properties.

Specifically, the Column Shapes Score was 97.86%, reflecting how well the distributions of individual variables were maintained in the synthetic data. The Column Pair Trends Score was 93.65%, demonstrating the preservation of relationships between different variables.

Individual column quality scores ranged from 95.00% for COMORBIDSUM to 99.75% for COMORB_HEART, with the majority of variables exceeding 97% quality. This high level of data fidelity enabled meaningful causal analysis while ensuring patient privacy, as no exact record matches were found between the synthetic and original datasets.

## Data Availability

All data produced in the present study are available upon reasonable request to the authors

## Notes

### Competing Interest Statement

The authors have declared no competing interest.

### Funding Statement

This study was partially funded by University of Calgary OpenDH program and Alberta Ministry of Mental Health

### Author Declarations

The study used (or will use) ONLY openly available human data that were originally located at Synthetic Data Vault which is an open-source ecosystem born in MIT Data to AI Lab

